# Development and validation of a multiplex bead based assay for the detection of antibodies directed against SARS-CoV-2 proteins

**DOI:** 10.1101/2020.09.02.20185199

**Authors:** Robert A. Bray, Jar-How Lee, Peter Brescia, Deepali Kumar, Thoa Nong, Remi Shih, E. Steve Woodle, Jonathan S. Maltzman, Howard M. Gebel

**Author notes:** **Corresponding Author:** Howard M. Gebel, Ph.D. Department of Pathology, Emory University Hospital, 1364 Clifton Rd. RM-HB-53, Atlanta, GA. 30322. Robert Bray and Jar-How Lee are co-first authors. Howard Gebel and Jonathan Maltzman are co-senior authors. ORCiDs Robert A. Bray, Ph.D.: 0000-0003-3598-5180. Jar-How Lee, Ph.D.: 0000-0002-2565-4276. Deepali Kumar, M.D.: 0000-0003-1961-0477. E.Steve Woodle, M.D.: 100000-0003-4280-0842. Jonathan S. Maltzman, M.D., Ph.D.: 0000-0001-5722-269X. Howard M. Gebel, Ph.D.: 0000-0003-0197-2752.

## Abstract

Transplant recipients who develop COVID-19 may be at increased risk for morbidity and mortality. Determining antibody status against SARS-CoV-2 in candidates and recipients will be important to understand the epidemiology and clinical course of COVID-19 infection in this population. There are multiple antibody tests to detect antibodies to SARS-CoV-2, but their performance varies according to their platforms and the antigenic targets, making interpretation of the results challenging. Additionally, currently available serological tests do not exclude the possibility that positive responses are due to cross reactive antibodies to community coronaviruses. This study describes the development and validation of a high throughput multiplex bead based antibody detection assay with the capacity to identify, simultaneously, patient responses to five distinct SARS-CoV-2 proteins. The antibody response to these proteins are SARS-CoV-2 specific as antibodies against four community coronaviruses do not cross-react. Assay configuration is essentially identical to the single antigen bead assays used in the majority of histocompatibility laboratories around the world and could easily be implemented into routine screening of transplant candidates and recipients. This new assay provides a novel tool to interrogate the spectrum of immune responses to SAR-CoV-2 and is uniquely suitable for use in the transplant setting.

## Introduction

Following the outbreak of the COVID-19 pandemic, assays using lateral flow, ELISA, chemiluminescence and fluorescence technologies were developed to detect antibodies to SARS-CoV-2 antigens (1-4). Target proteins include the SARS-CoV-2 nucleocapsid (NC) phosphoprotein (5), the full-length spike glycoprotein (6), or the receptor-binding domain (RBD) of the spike protein (7). The underlying premise is that such antibody assays can be used to evaluate the epidemiology of COVID-19 and, potentially, identify individuals and communities who would be protected from reinfection and have higher rates of herd immunity, respectively. However, it remains unclear which antibodies are actually protective or how long antibody levels are sustained (8). Notably, the majority of tests currently in use have reported sensitivities and specificities ranging from 96%-98%, which, while acceptable, are especially concerning when evaluating individuals from regions with <5% prevalence of disease (9, 10). Under those circumstances, a positive result has a high probability to be falsely positive, especially in a situation of low population prevalence. Since the majority of FDA approved serological assays only detect one viral target per test, even a true positive result has limited meaning, as only certain antibodies to SARS-CoV-2 appear to have viral neutralizing activity (11). Thus, simply considering a patient as SARS-CoV-2 antibody positive provides, at best, incomplete and at worst, misleading information regarding the clinical implications of those antibodies.

The limitations of the current assays to detect antibodies to SARS-CoV-2 become even more apparent when considering the spectrum of clinical responses to COVID-19. Clearly, certain patient characteristics are associated with an increased risk of morbidity and mortality (12). For example, recipients of solid organ allografts who become infected with SARS-CoV-2 are at especially high risk for adverse consequences compared to matched controls, presumably a consequence of being immunosuppressed and other underlying health issues (13). Knowing their SARS-CoV-2 antibody status before and after transplant could affect patient care.

Transplant candidates and recipients are routinely monitored to evaluate their HLA antibody status pre- and post-transplantation. The majority of histocompatibility laboratories around the world use a Luminex platform to detect and identify the HLA antibodies these patients possess (14). The HLA Luminex-based test is a multiplex, solid-phase antibody detection assay where up to 100 different microparticles coated with individual class I or class II HLA alleles, respectively, are simultaneously tested with patient sera. Due to its sensitivity, specificity and high throughput, this multiplex bead assay revolutionized the field of organ transplantation (15). In a similar manner, a Luminex-based multiplex test designed to identify antibody responses to unique SARS-CoV-2 targets could dramatically enhance our understanding of the immune response to COVID-19. In this study, we describe the development and validation of a multiplex high throughput solid phase assay that simultaneously determines the presence/absence of antibodies to five distinct SARS-CoV-2 viral proteins.

## Materials and Methods

### Target proteins

A prototype multiplex bead-based kit (LabScreen^TM^ COVID PLUS; Cat#: LSCOV01001) for the Luminex platform (Luminex, Corp., Austin TX) was developed by One Lambda Inc., Inc. (West Hill, CA). The assay includes four distinct fragments of SARS-CoV-2 Spike proteins, namely; 1) Full Spike extracellular domain; 2) Spike S1; 3) Spike, Receptor Binding Domain (RBD); and 4) Spike S2; The fifth target is the SARS-CoV-2 NC protein. Additionally, the kit incorporates Spike S1 fragments from six other coronaviruses, namely HCoV-229E, HCoV-HKU1, HCoV-NL63, HCoV-OC43, MERS-CoV and SARS-CoV. Proteins were obtained either from commercial sources namely, Sino Biologicals (Wayne, PA.), Euprotein, Inc. (North Brunswick, NJ.), RayBiotech (Peachtree Corners, GA.) or prepared in house (One Lambda Inc., Inc. West Hills, CA.). The source and catalogue numbers for each of these proteins is shown in supplemental table S1. The SARS-CoV-2 Full Spike, Spike S2 and SARS-CoV Spike S1were produced in-house based on published sequences (16,17)

### Polyclonal Antisera for protein validation

Polyclonal antisera, used to validate expression of the above proteins after their conjugation to individual beads. Antisera were purchased from a single commercial source, Sino Biologicals (Wayne, PA). The polyclonal antibodies used for bead validation were: SARS-CoV-2 NC Protein (Cat.#: 40588-T62, 1:1000 dilution), SARS-CoV-2 Spike (Cat#: 40589-T62; 1:1000 dilution), SARS-CoV-S2 (Cat#: 40590-T62; 1:1000 dilution), SARS-CoV S1 (Cat#: 40150-T62, 1:1000 dilution), SARS-CoV NC Protein (Cat#: 40143-T62, 1:1000 dilution), MERS-S1 (Cat#: 40069-T52, 1:1000 dilution), MERS NC Protein (Cat#: 40068-RP02, 1:1000 dilution), MERS Spike (CAT#: 40069-T30, 1:1000 dilution), MERS S2 (Cat#: 40070-T60, 1:1000 dilution) and HCoV-HKU1 S1 (CAT#: 40021-RP01, 1:1000 dilution).

### Patient Samples

Serum samples from both presumed negative patients, (i.e., pre-COVID-19), and confirmed COVID-19 positive patients were used in this study. Samples were provided by Emory University and the University of Washington. Initially, 96 serum samples from a pre-COVID-19 time period (October 2018) were randomly selected. Subsequently, serum from 42 convalescent patients with confirmed SARS-CoV-2 infection (none of them transplanted) were also selected for testing. SARS-Cov-2 infection was confirmed either by PCR testing (University of Washington or Emory University) or by a SARS-CoV-2 antibody testing by an internally developed and validated ELISA assay. Lastly, archived serum samples from 51 randomly selected candidates on the UNOS wait list in May 2020, were selected for testing. These samples were tested for SARS-CoV-2 RBD antibodies by ELISA.

### SARS-CoV-2 antibody detection in patient sera

Antibody detection on antigen-coated microparticles was performed as follows: Five microliters of viral antigen coated beads were admixed with 20 μl of a 1:10 dilution, in PBS, of patient or control serum that was pre-treated with 10mM EDTA. The serum/bead admixture was incubated in the dark at 20–25°C for 30 minutes with gentle rotation followed by three sequential washes, each using 200μl wash buffer (WB) (OLI Cat. # LSPWABUF). Following incubation, 100µl of a pre-titered PE-conjugated anti-human IgG (Jackson ImmunoResearch, West Grove, PA.: CAT#: 109-116-170) was added. Next, the beads were vortexed and incubated, in the dark, for 30 minutes at 20 - 25º C with gentle shaking. Finally, the beads were washed twice with 200μl WB. Microparticles were resuspended in 80μl of 1X PBS and analyzed on a Luminex FLEXMAP 3D^®^ instrument (Luminex Corp. Austin, Tx.). Final analysis of MFI (Mean Fluorescence Intensity) was performed using Microsoft Excel.

### Calculation of cut-off, sensitivity, specificity and accuracy

To determine the positive/negative cut-off values for each recombinant protein, we used the Mean +3xSD formula based on the mean MFI values of 96 COVID-19 negative samples collected prior to 2019. The 3xSD result obtained was considered as the cut-off value for each antigen.

## Results

### Validation of SARS-CoV2 coated microparticles

Microparticles coated with the SARS-CoV-2 proteins, Full Spike, S1, S2, RBD and NC as well as non-SARS-CoV-2 proteins from community coronaviruses (CoV-229E-S1, HCoV-HKU1-S1, HCoV-NL63-S1, HCoV-OC43-S1), and novel coronaviruses (MERS-S1, and SARS-S1), were evaluated with commercial polyclonal antibodies in a matrix fashion as shown in Figures 1A and 1B. For the SARS-CoV-2 proteins (Figure 1A), antisera specific for the NC showed positive reactivity with the NC-coated bead while failing to react with the other 4 SARS-CoV-2 proteins. The antisera directed against the Full Spike protein reacted with the Full Spike, S1 and RBD coated beads and was non-reactive with the S2 and NC. This reactivity pattern was in agreement with the manufactures published specifications for this antibody. The antisera directed against the SARS-CoV-2 S2 protein showed positive reactivity against both the Full Spike and S2 proteins as anticipated based on the manufacturers published specifications for this antibody. The antisera specific for the SARS-CoV-2 Spike protein showed cross-reactivity against SARS-CoV-2 Full Spike, S1 and RBD. The antisera specific for SARS NC protein also showed cross-reactivity with the SARS-CoV-2 NC protein consistent with the published specifications for this antibody.

**Figure 1.**
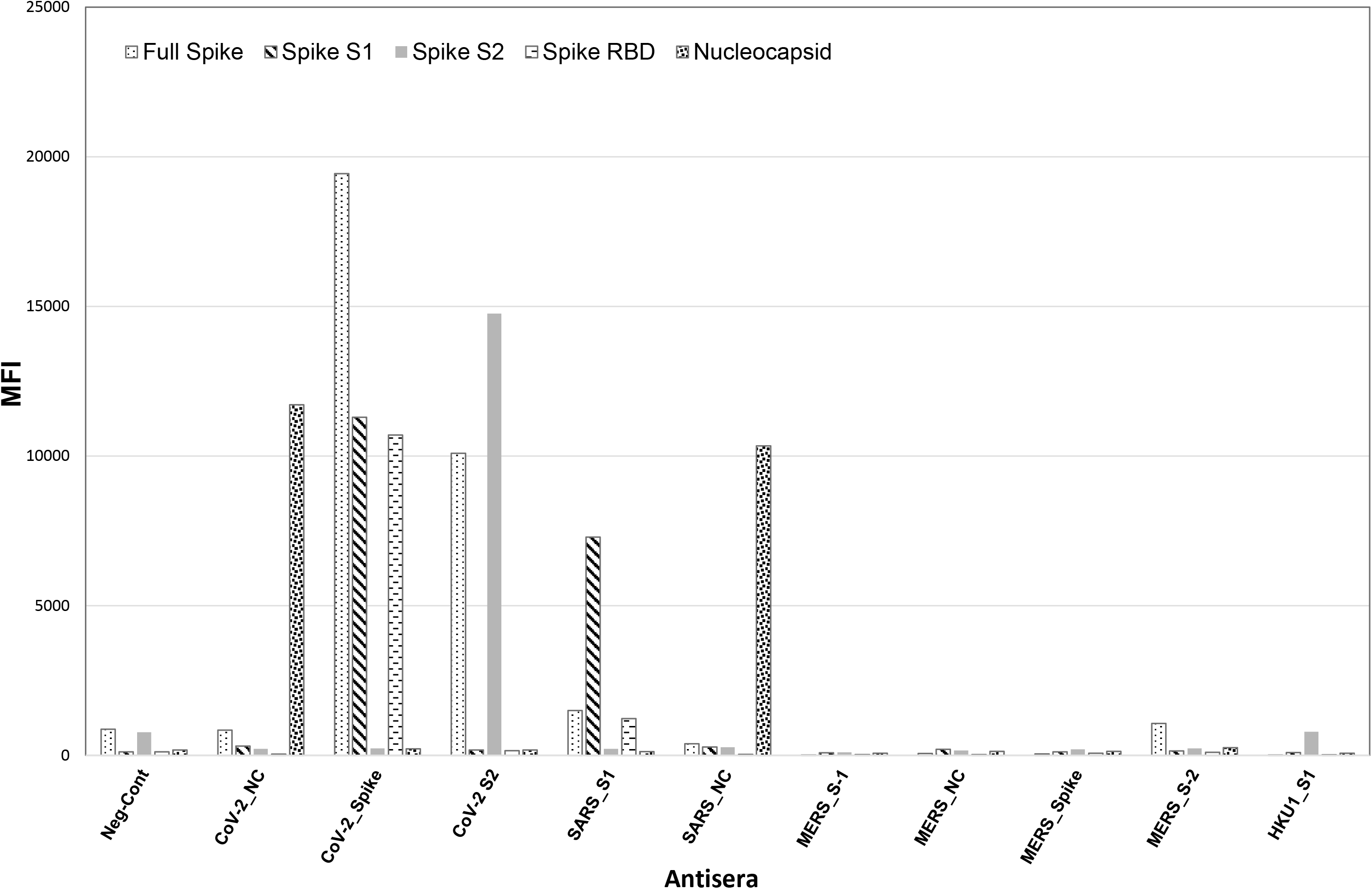
SARS-CoV-2 and non-CoV-2 proteins were tested with a battery of allo-antisera. Reactivity of the SARS-CoV-2 proteins with antisera reactive with SARS-CoV-2 nucleocapsid, SARS-CoV-2 full spike protein, SARS-CoV-2 S2 protein as well as other non-SARS-CoV-2 antisera are shown. The x-axis represents the allosera used and the y-axis the MFI value of the reactivity.

### Pre-COVID-19 samples vs SARS-CoV-2 proteins

Figure 2A shows data from a representative experiment wherein serum samples, collected from 96 randomly selected and pre-COVID-19 (October, 2018) kidney transplant candidates were tested with the COVID Plus assay. As the data illustrate, with the exception of the S1 spike protein, all samples had MFI levels ≤5000. For the Full Spike protein, the average value was 675 + 816; range 44 – 4540 MFI. For the spike S2 protein, the average value was 138 + 97; range 42 – 908 MFI. For the spike RBD protein, the average value was 224 + 363; range 23 – 3000 MFI. For the NC protein, the average value was 388 + 441; range 89 – 2593 MFI. For the S1 protein, background levels were higher but all samples showed MFI values <7,500. For the S1 spike, the average background value was 1226 + 1411; range 133 to 7439 MFI. Based on these data, responses <5000 MFI for the Full Spike, S2, RBD and NC coated beads were considered negative, while values <7500 MFI were considered negative for S1. Figure 2B displays the responses for the 96 pre-COVID-19 samples against a panel of other coronavirus targets including SARS-CoV and MERS. The data demonstrate that while most individuals have significant IgG responses to community coronaviruses, all pre-COVID-19 samples tested negative against the SARS-CoV-2 targets as well as to SARS and MERS S1 proteins.

**Figure 2.**
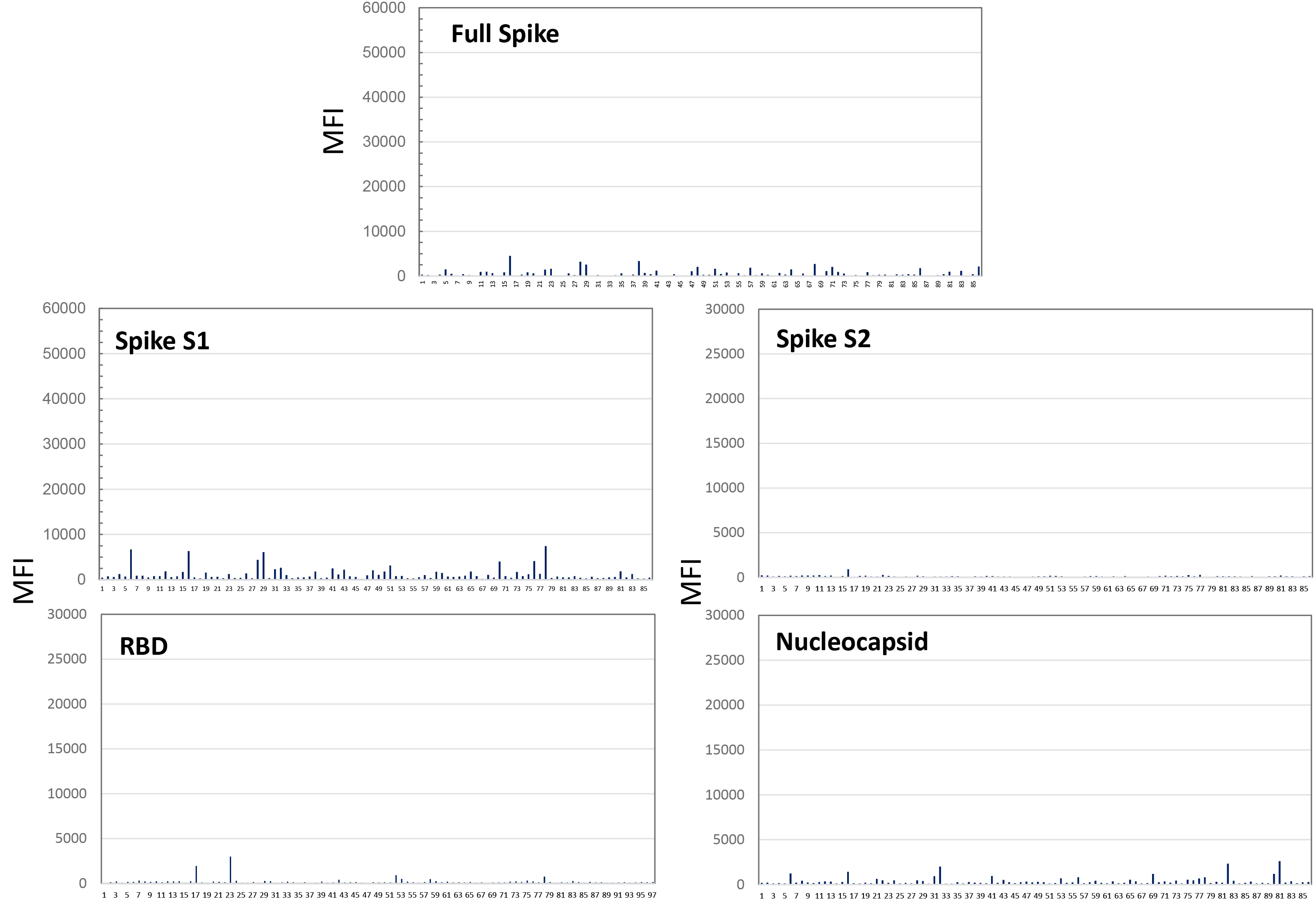
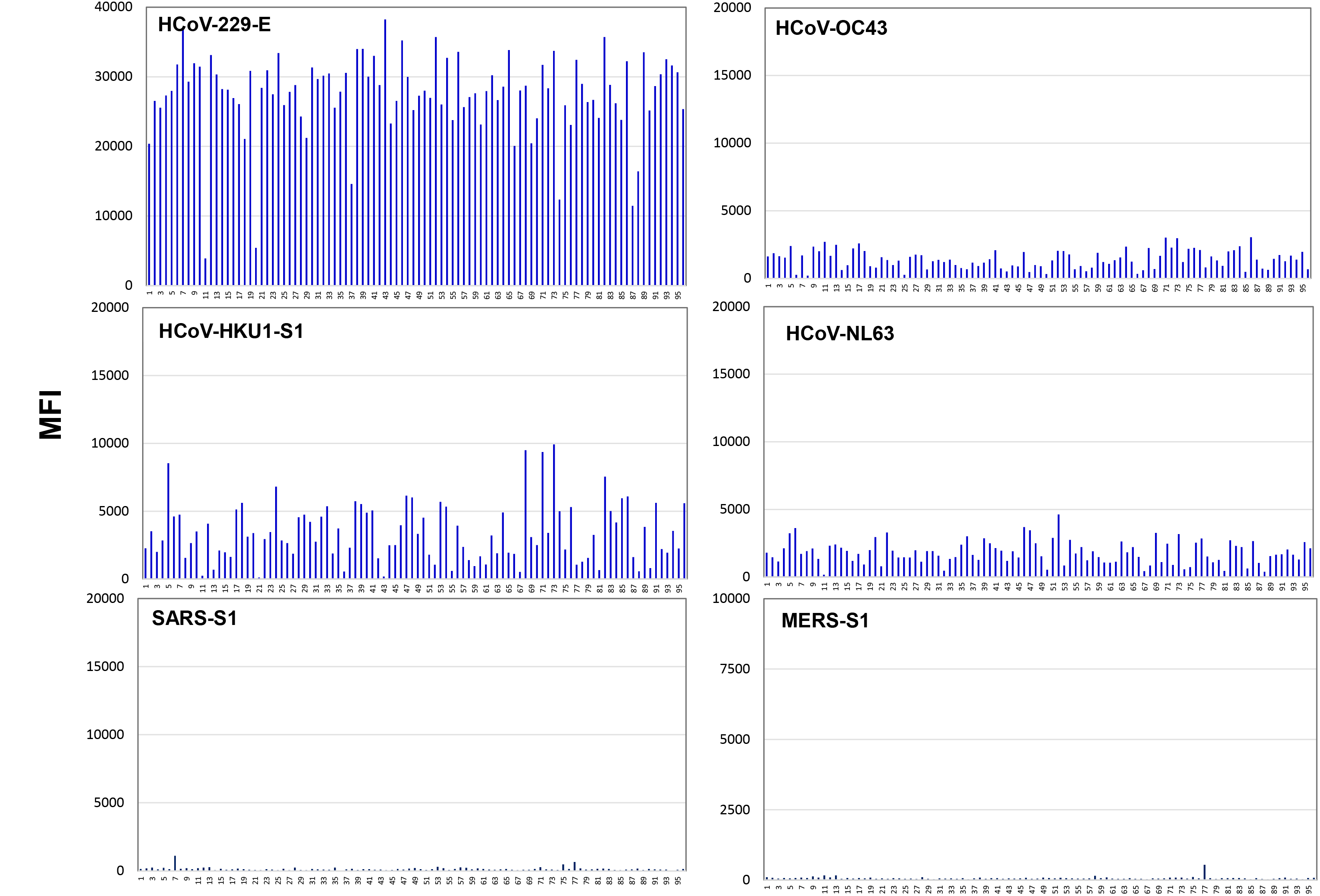
Reactivity of 96, pre-Covid-19, serum samples from candidates on the kidney wait list tested against the 5 SARS-CoV-2 proteins in the assay. Samples used were from October 2018. The x-axis represents the sample number and the y-axis the MFI value of the reactivity. **2A**. Shows reactivity against the panel of SARS-CoV-2 proteins. **2B**. Shows reactivity against a panel of other corona viruses associated with similar upper respiratory infections including the common cold.

### SARS-CoV-2 positive samples vs SARS-CoV-2 proteins

Sera from 42 non-transplanted patients each confirmed positive for SARS-CoV-2 infection by either RT-PCR positivity or SARS-CoV-2 antibody testing (ELISA, Emory University) (18) were tested with the COVID-Plus assay. As shown in Figure 3, serum from all 42 patients tested positive with at least three of the five SARS-CoV-2 proteins. Positivity was determined as an MFI value >3 standard deviations above the mean of the negative controls. Interestingly, the level of responses differed from patient to patient. Specifically, MFI values ranged from 10,000 - 60,000 MFI for the Full Spike protein, 5,000 - 50,000 MFI for S1, 2,000 – 8,000 for S2, 2,000 - 30,000 for RBD and 3,000 - 15,000 for the NC protein. All 42 patients had a >5,000 MFI response to the Full Spike, while 33/42, 38/42, and 39/42 patients had >5,000 MFI response to S2, RBD and NC, respectively. For S1, 38/42 samples showed a response >7,500 MFI.

**Figure 3.**
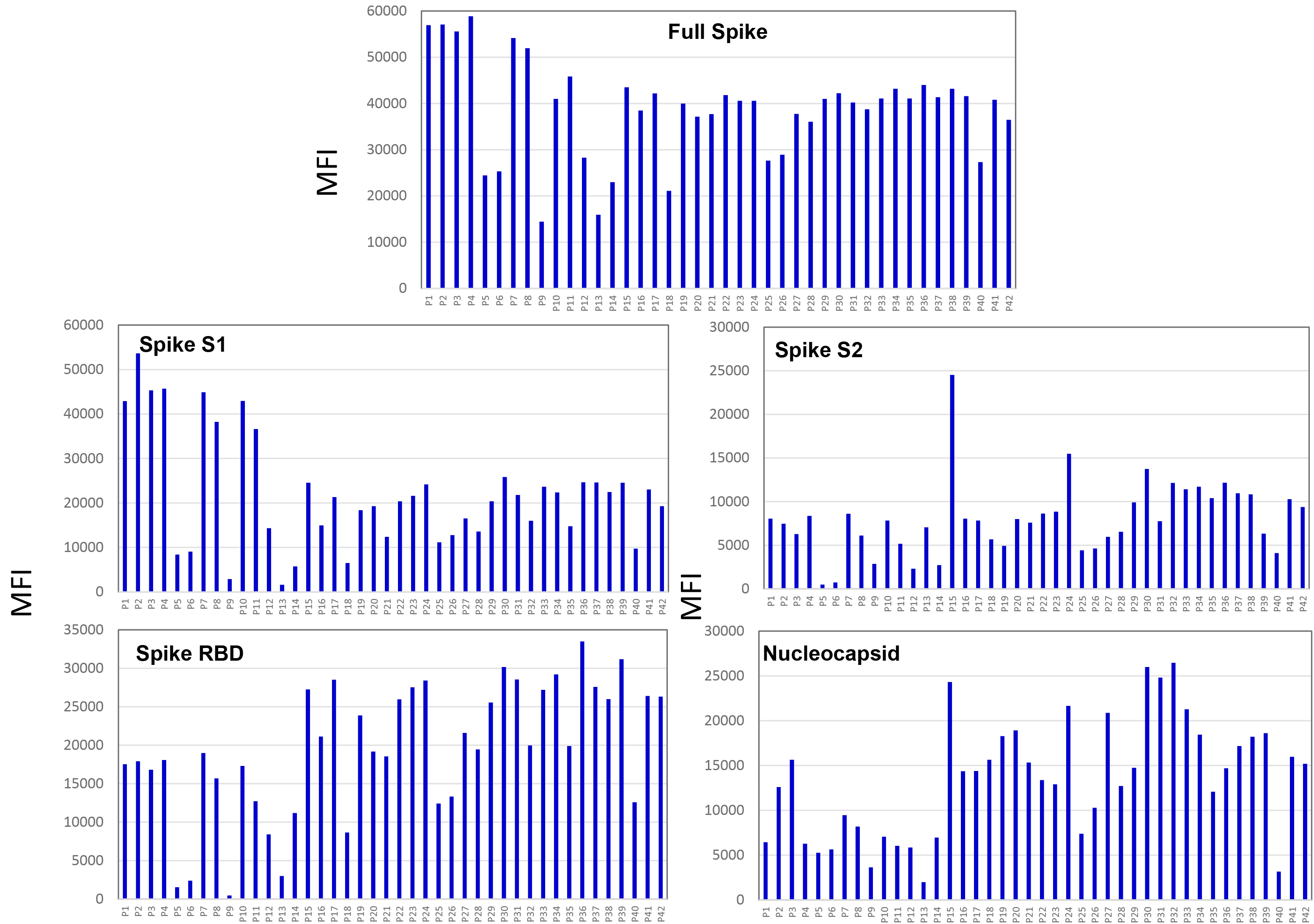
Reactivity of 42 SARS-COV-2 confirmed positive individuals against the panel of 5 SARS-CoV-2 proteins. The x-axis represents the sample number and the y-axis the MFI value of the reactivity.

### Differential responsiveness of RT-PCR positive patients

The spectrum of response to SARS-CoV-2 proteins is illustrated in Figure 4. In figure 4A, the semi-quantitative MFI differences between and within individuals is plotted. Figure 4B illustrates the proportional difference among the COVID-19 positive patients. The data show that, on average, the Full Spike protein constituted the majority of the response in any given individual. However, the proportional differences between the three components of the Full Spike S1, S2 and RBD varied greatly between individuals. Of note, the proportional response to the RBD protein varied from 2% to 26% of the total MFI response. For cumulative MFI values <40,000, the RBD response ranged between 2 and 10% with a mean response proportion of 5.5%. However, for patients with a cumulative MFI value >40,000, the RBD response ranged between 12-26% with an average of 20%.

**Figure 4.**
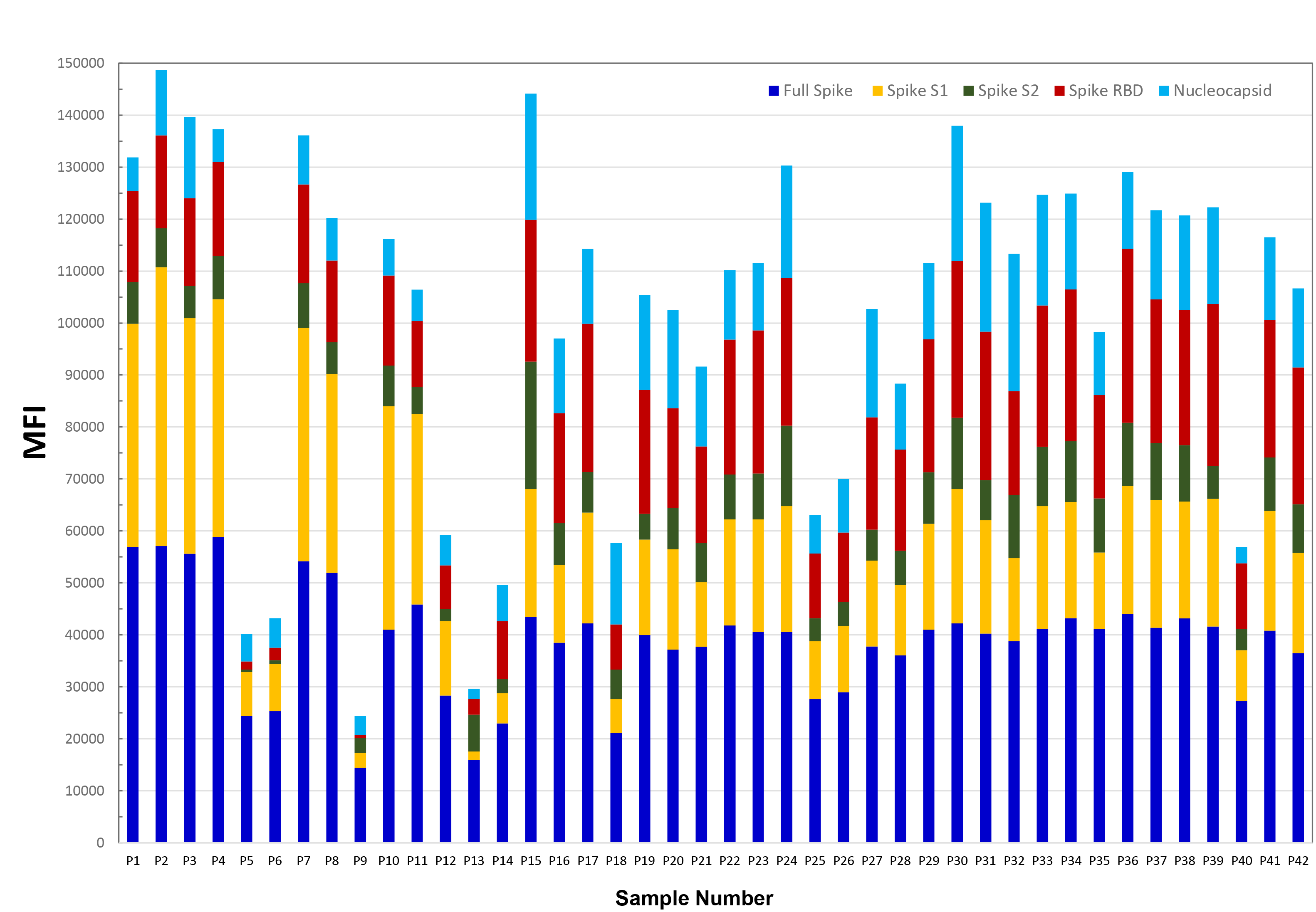
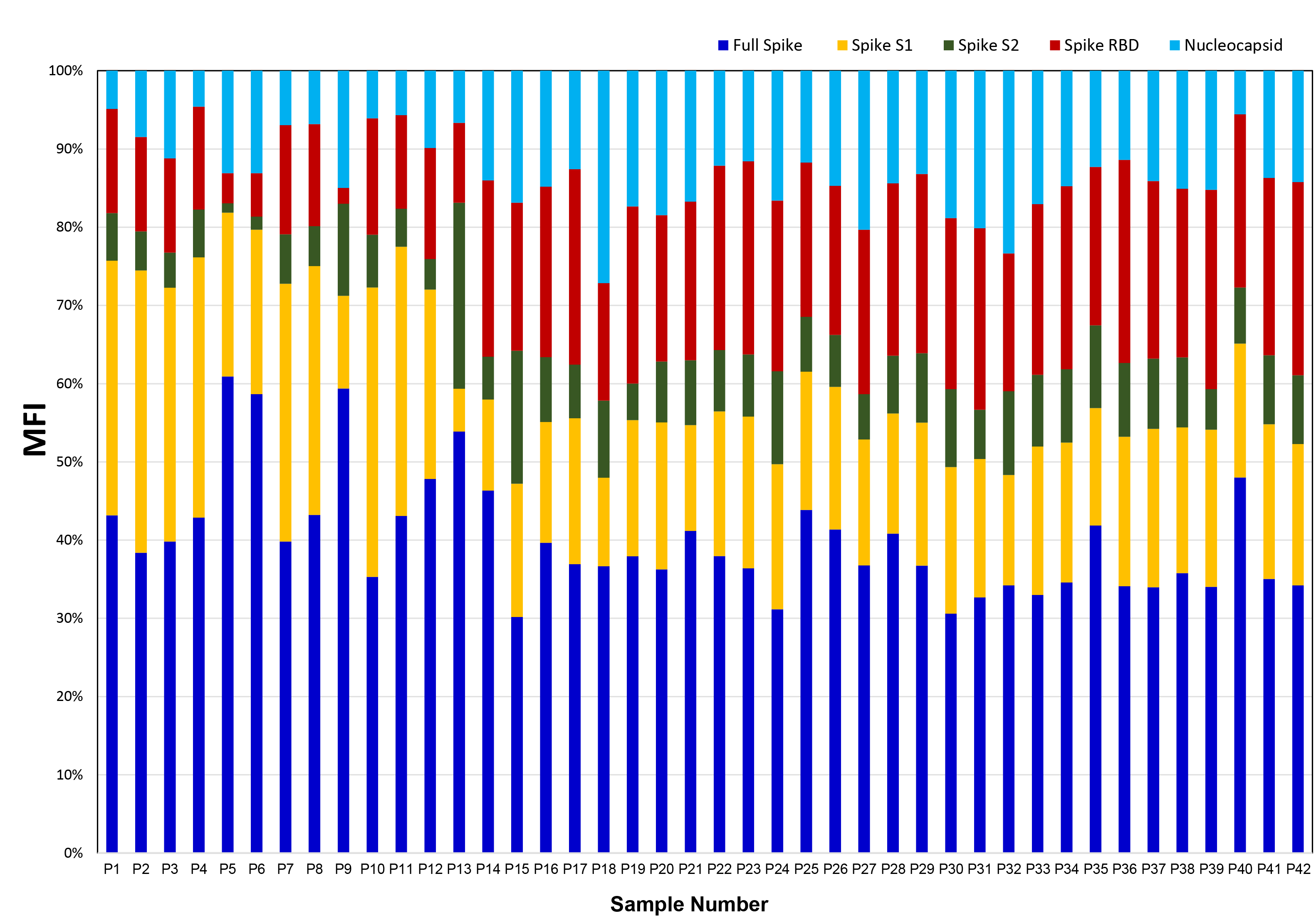
Reactivity patterns of the 42 confirmed SARS-CoV-2 positive patients. Figure **4A**. The height of each histogram bar denotes the cumulative MFI value for all tested proteins for each patient. The various colored sections within each histogram bar illustrate the MFI contribution, to the total MIF, for each individual protein. Figure **4B**. Histogram plot of each patient showing the proportional contribution, to the total cumulative MIF value, of each component protein. This plot demonstrates the proportional variability of individual protein responses between individuals.

### Screening of serum samples from transplant candidates collected post-COVID-19 onset

As seen in Figure 5, 3/51 serum samples collected in May, 2020 from renal transplant candidates tested positive for at least one SARS-CoV-2 protein. Serum from two patients reacted with all five proteins (subjects #14 and #51), while one subject (#8) reacted only with the Full Spike protein. Interestingly, these same samples were tested for RBD antibodies by ELISA methodology. While patients 14 and 51 were positive, patient 8 was negative. To better understand these Luminex positive reactions, historic sera from these three subjects, dating back to pre-CoVID-19 time periods, were tested for reactivity in the COVID-Plus assay. Subjects #14 and #51 initially showed a distinct pattern of negative reactivity until May and April of 2020, respectively (Figure 6A and 6B). The positive results in April and May are consistent with COVID-19 infection during a period of relatively high virus prevalence in Georgia. In contrast, subject #8 showed reactivity only against the Full Spike protein and this reactivity appeared long before the global appearance of COVID-19 (Figure 7).

**Figure 5.**
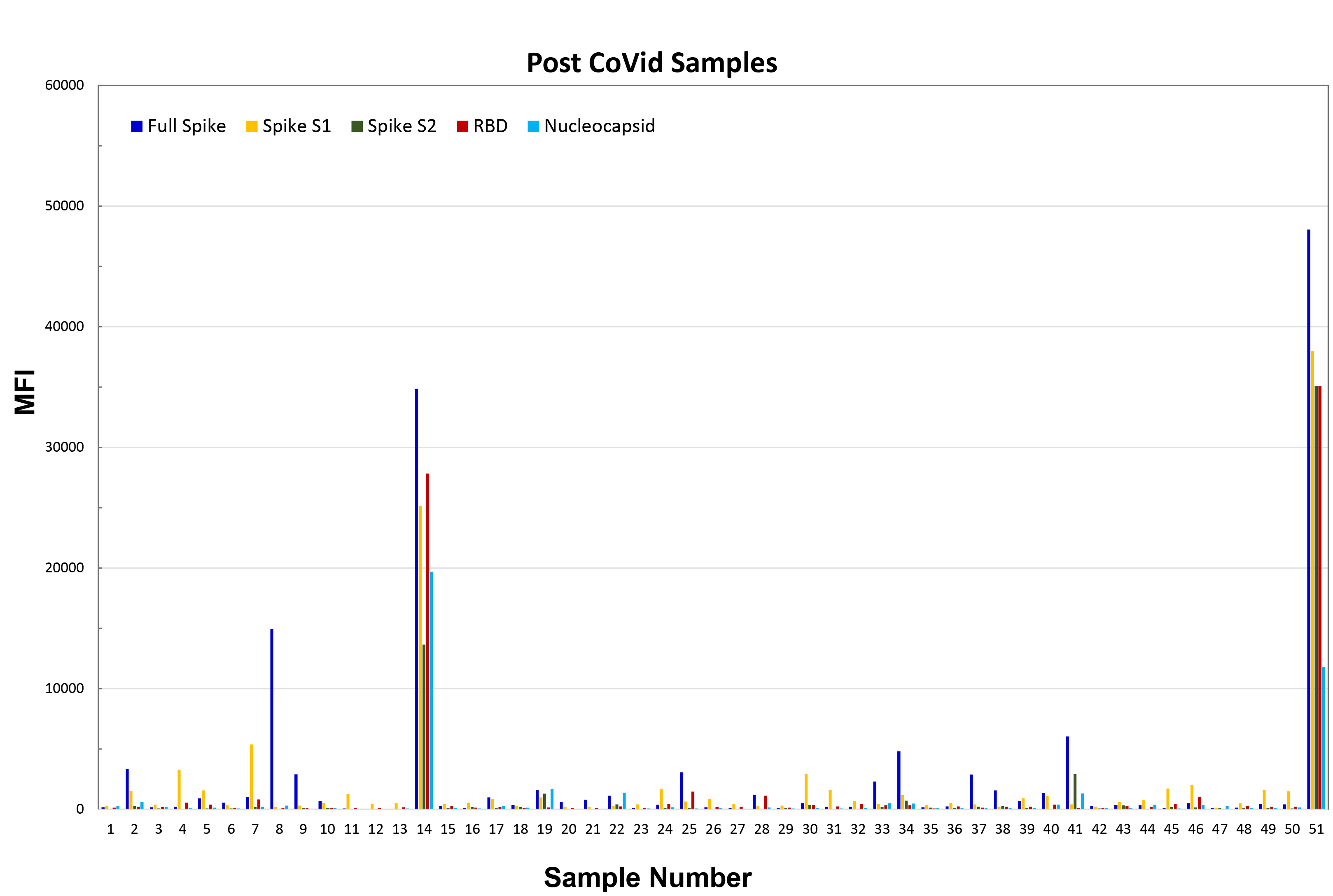
Reactivity of 51, post Covid-19 samples against the COVID-PLUS panel. Samples were obtained from candidates on the kidney wait list during the month of May 2020. Note that patients #14 and #51 would be scored as positive for antibody. Patient #8 represents an apparent false positive with reactivity against only the Full Spike protein.

**Figure 6.**
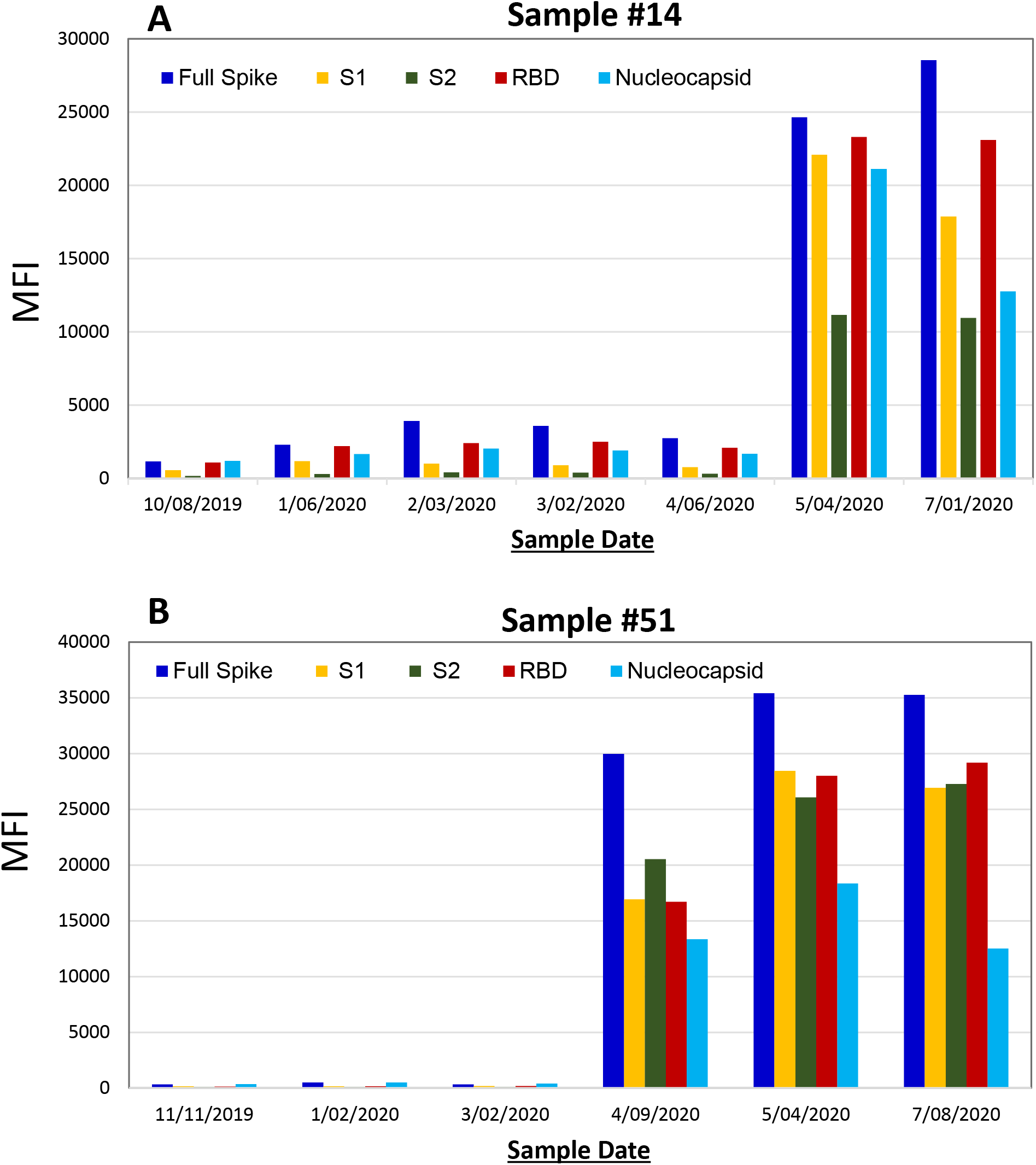
Chronological testing serum samples from patients 14 and 51 for SARS-CoV-2 antibodies. The x-axis represents the sample date and the y-axis the MFI value of the reactivity.

**Figure 7.**
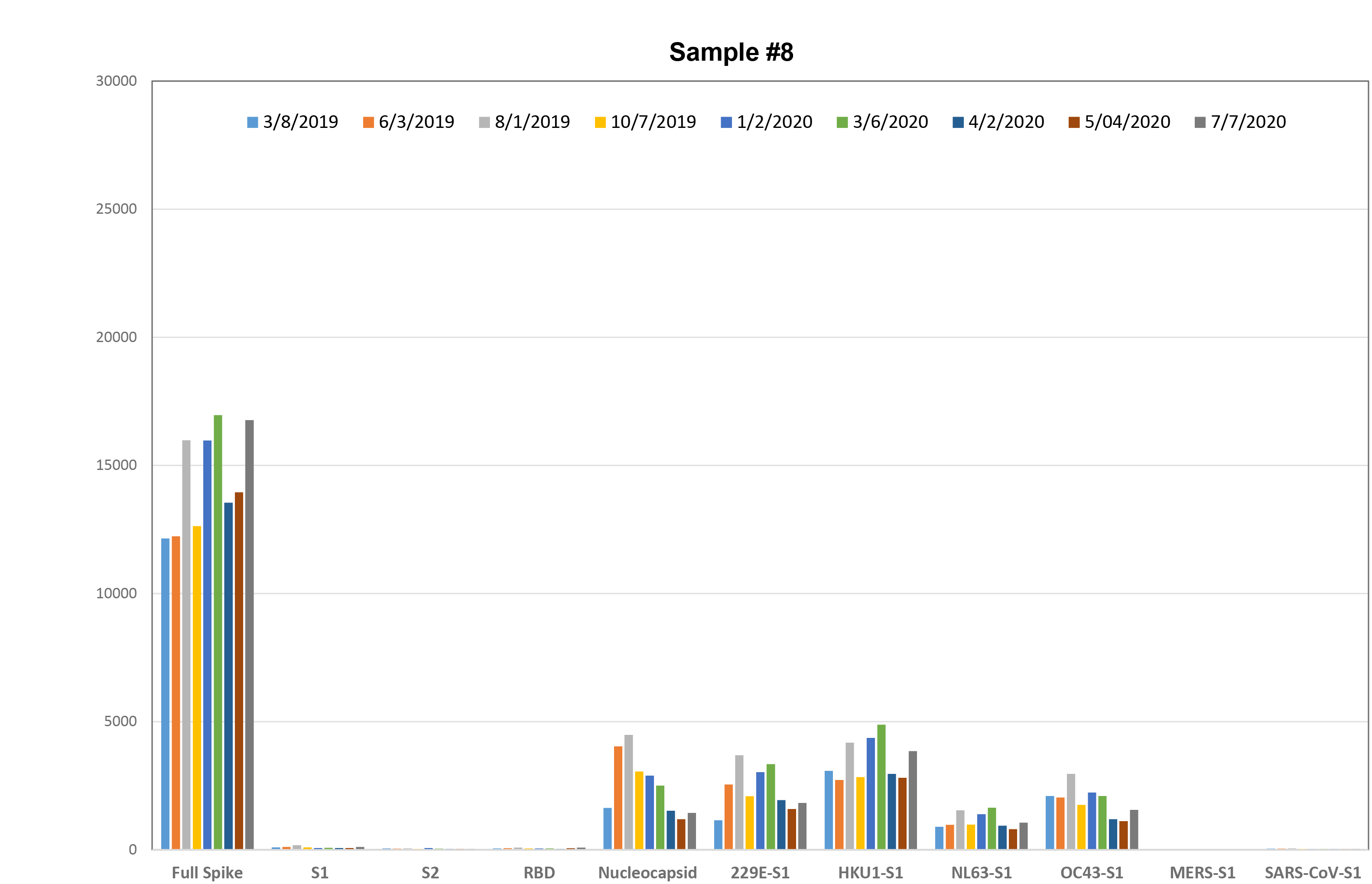
Chronological graph of serum dates from transplant candidate #8 showing presence of likely false positive reactivity dating back to a pre-COVID-19 era. The x-axis represents the sample date and the y-axis the MFI value of the reactivity.

## Discussion

Herein, we describe the development of a novel multiplexed Luminex based immunoassay that simultaneously and semi-quantitatively assesses patient sera for antibodies to multiple SARS-CoV-2 proteins. This assay builds on >25 years of experience with the Luminex platform to detect and identify antibodies to HLA antigens present in the serum of transplant candidates and recipients. The strengths of this multiplexed, solid phase assay compared to other platforms (e.g., ELISA) include the ability to assay multiple viral targets simultaneously (and thereby provide internal assay controls for coronavirus specificity), the capacity to incorporate additional SARS-CoV-2 proteins in the future, a relatively short assay time, high throughput and semi-quantitative assessment of the antibody response. Furthermore, these data demonstrate that the antibody responses to common community coronaviruses do not cross react with the SARS-CoV-2 proteins in the COVID Plus assay (Figure 2). An additional benefit is that evaluation and monitoring of samples from transplant candidates and recipients for antibodies to SARS-CoV-2 is readily adaptable into the routine clinical practice of histocompatibility laboratories that support solid organ and/or stem cell transplant programs.

In the United States, the need for robust, accurate and reproducible serological tests to identify individuals who developed antibodies to SARS-CoV-2 was recognized by mid-March 2020. In response to demand, the FDA authorized emergency use authorization of a multitude of such tests to be developed, implemented and used throughout the US. It was anticipated that the tests would identify individuals 1) who had developed humoral immunity in response to exposure; 2) would be resistant to reinfection with SARS-CoV-2 and 3) from whom convalescent plasma could be collected and used as a therapeutic or prophylactic for patients diagnosed with COVID-19 (19). However, recent studies reveal that a positive antibody test is not necessarily adequate to determine current or future immunity to SARS CoV-2. Indeed, recent data by Sutar et al (18) documented the importance of appropriate timing for serological testing relative to PCR testing and/or symptom onset after infection. Additional studies by Chi et al revealed that although antibodies to the RBD of SARS CoV-2 are the antibodies most likely to be neutralizing, so, too, could antibodies to other viral targets (20). Thus, understanding which antibodies are protective and being able to detect them are both essential components of serological testing.

The genome of SARS-CoV-2, a single stranded, enveloped RNA coronavirus, encodes four major structural proteins, namely, spike, envelope, membrane and nucleocapsid as well as more than a dozen non-structural proteins (21). Based on their immunogenicity and predicted neutralizing potential, either the NC, spike (full length, S1 or S2) or the RBD are the individual designated targets for the vast majority of antibody assays in use. Sensitivity and specificity for such assays typically ranges from 96-98%, which, while reasonable, can unfortunately lead to false positive and false negative results (9,10). An additional important limitation of such single target assays is that they fail to assess the breadth of a patient’s response. In fact, recent studies reveal that patient antibody responses to SARS CoV-2 are not uniform. For example, while convalescent plasma donors and patients who have recovered from SARS-CoV-2 infection have detectable RBD antibodies that are neutralizing, RBD antibodies detected in pediatric COVID-19 patients who developed multisystem inflammatory syndrome are not neutralizing (11).

The collective published data illustrate the gap in our understanding the complexities of the humoral immune response to COVID-19. Assays that interrogate a response to a single viral antigen limit our ability to understand fully the immune responsiveness to SARS-CoV-2. This is evidenced by the response of subject #8 who showed strong reactivity against the Full Spike protein but was negative for S1, S2 and RBD. The low level of NC protein reactivity also did not exceed the 3 SD cutoff to consider the reaction as positive. Since S1, S2 and RBD proteins are elements of the Full Spike protein; this reactivity could reflect responsiveness to a unique cross-reactive epitope not related to SARS-CoV-2 infection. This reactivity pattern was not observed in any other SARS-CoV-2 negative sample either pre- or post-COVID-19. Furthermore, the reactivity pattern of subject #8 extended well back into the global pre-COVID-19 era. The exact nature of this reactivity is under further investigation. When the results of all confirmed SARS-CoV-2 positive samples and negative samples obtained from samples prior to the COVID-19 pandemic are considered, the specificity and sensitivity of the assay is 98.6% and 100%, respectively. Excluding the data from the one subject positive only with the Full Spike protein, assay specificity and sensitivity are both 100%.

One recently described assay simultaneously evaluated the response to both nucleocapsid and spike proteins with a reported sensitivity of 100% and 99.9% sensitivity (22), However, that assay required patient samples to be diluted 1/200 before testing, which suggests that the signal to noise ratio of undiluted samples was less than optimal. Furthermore, the assay time was close to 20 hrs. In the multiplex assay described here, the responses to five different viral targets were simultaneously tested and the total assay time was <4 hours. Furthermore, 96 samples can be assessed at the same time in a single tray.

Interestingly, the proportional responses to the five antigens differed among the patients. For example, some individuals displayed a higher MFI to NC vs RBD (e.g., sample P18; 15,641 vs 8662, respectively), while other subjects displayed a higher response to the RBD target than to the NC (e.g., sample P36; 33,491 vs 14,703, respectively). It is intriguing to speculate whether such observations would explain differences in clinical manifestations and outcomes. The differences could potentially be derived from immune deviation because of underlying disease or genetics. There may also be influences due to inter-individual variations in B cell repertoires, differences in disease progression, and the amount of time from which patients were initially diagnosed until the time they were tested for antibodies. Interrogation of antibody responses from well-characterized patients grouped according to age, severity of disease, and response to therapy will be improved by simultaneously evaluating the response to multiple SARS-CoV-2 proteins. Other important questions such as an individual’s antibody response to vaccination, viral neutralization titers and whether some antibodies enhance disease progression will be better addressed with this technology.

The semi-quantitative nature of the response offered by this assay will also likely be of greater value rather than the simple “yes” or “no” result obtained from lateral flow assays. The titer of antibodies to SARS-CoV-2 correlates with their ability to limit viral infectivity. Our preliminary data suggest that due to the increased dynamic range of the Luminex FLEXMAP 3D^®^ instrument, multiplex testing for SARS-CoV-2 antibodies allows for a level of quantification not possible with lateral flow, ELISA or chemiluminescence assays unless serial dilutions are performed for each sample.

While this first iteration of this multiplex Luminex based assay has five target proteins, our intention is to add additional targets to the panel. Indeed, the SARS-CoV-2 virus has 29 total proteins any of which could be immunogenic and stimulate an antibody response. Antibodies to non-structural and accessory proteins of SARS-CoV-2, namely ORF9b and NSP5, have been identified in the convalescent sera of infected patients infected with SARS-CoV-2 (23). Although viral targets not expressed on the cell surface are not typically considered to generate antibodies that are neutralizing, that is not always true. For example, for influenza, antibodies to ORF proteins confer protective immunity based on passive transfer studies and experiments with B cell knockout mice (24, 25). As the current version of COVID Plus only uses a total of thirteen individual Luminex addresses, there is ample real estate for expansion.

From a more practical standpoint, the SARS-CoV-2 multiplex assay described here is very similar to the multiplex assays used to identify HLA antibodies routinely deployed by HLA laboratories around the world. As such, incorporating the SARS-CoV-2 assay into routine testing of transplant candidates and recipients would be seamless, and provide useful information to the surgeons and clinicians caring for patients during the COVID-19 pandemic.

Limitations of this study include the source of our negative and positive control sera. Negative control samples were obtained from patients with chronic kidney disease. As such, the patients may be limited in their ability to produce antibodies. However, we observed that all the sera from this group of subjects had antibodies to community CoV and, furthermore, sera from two patients that had tested negative prior to the COVID-19 pandemic did develop antibodies to all five SARS-CoV-2 at a time infections were surging. With regard to sera from confirmed positive cases, we were unable to collate sample date with disease status (e.g., acute vs convalescent). A final limitation to the multiplex assay (and all other antibody detection assays) is that it does not determine whether any of the detected antibodies have neutralizing capacity. Future studies will address this issue.

In conclusion, we developed a multiplex, high throughput, sensitive and specific assay that should provide a more comprehensive and in-depth understanding of the dynamics, biology and repertoire of antibody responses to SARS-CoV-2. This assay will be easily adaptable in the transplant setting.

## Data Availability

Data available upon reasonable request.

## Acknowledgements

The authors would like to acknowledge Drs. Sean Stowell, Scott Boyd and Zahra Kashi for testing and/or providing COVID-19 positive samples. The authors also thank Ms. Shilpee Biwas for expert technical assistance and Dr. Tina Meng for Spike protein preparations. In addition, the authors acknowledge Drs. Medhat Askar and Phillip Ruiz for helpful discussions regarding assay development.

## AUTHORS’ CONTRIBUTIONS

RAB: designed the study, collected the data, analyzed the data, interpreted the data, drafted the article and revised the article critically.

J-H Lee: designed the study, collected the data, analyzed the data, interpreted the data, drafted the article and revised the article critically.

PB: collected and analyzed the data.

DK: designed the study, interpreted the data and revised the article critically

TN: collected and analyzed the data.

RS: collected and analyzed the data.

ESW: designed the study, interpreted the data and revised the article critically.

JSM: designed the study, collected the data, analyzed the data, interpreted the data, drafted the article and revised the article critically.

HMG: designed the study, collected the data, analyzed the data, interpreted the data, drafted the article and revised the article critically.

**Abbreviations**
NC: SARS-CoV-2 Nucleocapsid Protein
S1: SARS-CoV-2 Spike S1 Protein
S2: SARS-CoV-2 Spike S2 Protein
RBD: SARS-CoV-2 Receptor Binding Domain
MFI: Mean Fluorescence Intensity
ELISA: Enzyme-Linked Immunosorbent Assay
HLA: Human Leukocyte Antigen

